# Explainable drug side effect prediction via biologically informed graph neural network

**DOI:** 10.1101/2023.05.26.23290615

**Authors:** Tongtong Huang, Ko-Hong Lin, Rodrigo Machado-Vieira, Jair C Soares, Xiaoqian Jiang, Yejin Kim

## Abstract

Early detection of potential side effects (SE) is a critical and challenging task for drug discovery and patient care. In-vitro or in-vivo approach to detect potential SEs is not scalable for many drug candidates during the preclinical stage. Recent advances in explainable machine learning may facilitate detecting potential SEs of new drugs before market release and elucidating the critical mechanism of biological actions. Here, we leverage multi-modal interactions among molecules to develop a biologically informed graph-based SE prediction model, called HHAN-DSI. HHAN-DSI predicted frequent and even uncommon SEs of the unseen drug with higher or comparable accuracy against benchmark methods. When applying HHAN-DSI to the central nervous system, the organs with the largest number of SEs, the model revealed diverse psychiatric medications’ previously unknown but probable SEs, together with the potential mechanisms of actions through a network of genes, biological functions, drugs, and SEs.

## 1. Introduction

The side effect (SE) remains a major challenge in patient care and brings fundamental costs to drug development^1^. Detecting SEs is a critical part of developing drugs and treating patients since SEs can cause significant morbidity and mortality^2, 3^. Unforeseen SEs lead to drug withdrawal during development—roughly six hundred drugs have been discontinued during trials or withdrawn from post-marketing surveillance^4, 5^. According to statistics from the FDA Adverse Events Report Systems (FAERS)^6^, the number of SE reports reached 2.3 million and led to 190,000 deaths in 2021. Among them, 1.4 million cases reported at least one unforeseen SE that was not currently described in product labeling. Predicting SEs in the early stages of drug development is thus critical to prevent the devastating consequences of SEs. Although detecting SEs by in-vitro or in-vivo models can provide preclinical evidence on SEs with underlying biological mechanisms, it is not cost-effective for many drug candidates^7^.

Furthermore, relying on clinical trials to detect SEs may face several challenges, and indeed, numerous SEs have been only observed after market release^8, 9^. This is because vulnerable populations were not well represented in trial participants, and the drugs were given for shorter durations in clinical trials than in actual use^10^.

To address these challenges, in-silico prediction of SEs, particularly using machine learning, has gained attention as identifying potential SEs that might occur in populations by utilizing large-scale molecular data. Neighboring methods have utilized similarity among drugs to extrapolate possible SEs for drugs of interest^11–13^. They construct drug neighbors using the *k*-nearest neighbors (KNN) algorithm. They assume that similar drugs might cause the same SEs because they share common chemical, biological, or phenotypic information. Furthermore, matrix decomposition methods^14–17^ extract latent features of both drugs and SEs. They characterize drugs via their chemical structures or target genes, and characterize SEs via associated genes^16^, associated drugs^18^, or SE categories^19, 20^. These conventional SE prediction methodologies often derive similarity using one type of interactions (single modality). Underlying mechanisms of SEs, however, comprise multimodal interactions among a variety of biological entities within an organism, aforementioned methods have limitations in incorporating such multimodal interactions.

To overcome this limitation and better represent the multimodal interactions around SEs, we utilize the knowledge graph, a data structure with heterogeneous relations and entities to represent multimodal interactions among genes, gene ontology, drugs, and SEs. Graph machine learning approaches can quantify the topology of graphs. With an advance in representation learning based on deep neural networks, graph neural network (GNN) has gained attention thanks to its flexibility in converting graph structure and entity features into condensed representation. GNN models^21^ use message aggregation and passing approaches to incorporate features from order-free neighbor nodes. A variety of GNN-based SE prediction models have been introduced ^22–24^, particularly for polypharmacy effects, using drug-gene^25, 26^ and gene-gene^27^ interactions. These studies leverage common gene features between two drugs to estimate edge types representing polypharmacy effects. However, they made limited efforts in the explainability of their individual SE prediction. The explainability of the SE prediction is very crucial because the molecular interactions that contribute to the SE prediction can elucidate the pharmacodynamics, which is sometimes unknown^28, 29^.

There are several studies investigating how to use machine learning approaches to explain biological mechanisms. Biologically informed deep learning has shown promising explainability in previous studies^30, 31^. Elmarakeby et al.^30^ designed biologically informed paths to constrain the connection between neurons in the fully-connected neural network. They derived meaningful paths from the importance score during backpropagation, whereas the fully-connected network could not explain the mechanism behind the interactions between two entities. Ghosal et al.^31^ presented important pathways of hierarchical biological processes by using an attention-based graph neural network. But they only considered the relation between biological processes, and thus failed to explain the impact of multimodal interactions. More importantly, the previous strategy is heavily dependent on the quality of hardcoded biological priors (gene ontology, pathways, etc.), which are sometimes incomplete and erroneous due to the complexity and heterogeneity of biological mechanisms. To address the limitation, we propose to infuse biological knowledge at the "semantic" level by interaction modality. This middle ground between the hardcoded priors and no priors can allow broad yet plausible boundaries and guide information to cascade through the boundary. Therefore, we propose an end-to-end **Hierarchical Heterogeneous Graph Attention Network (HHAN) model for drug-SE interaction (DSI) prediction**, namely **HHAN-DSI**. Our novel model constrains the information propagation and node aggregation only for biologically guided paths^32^ and hierarchy. Further, such biologically guided GNN learning also allows us to explain the mechanism of action between drugs and SEs.

## 2. Results

### 2.1 HHAN-DSI model development

#### 2.1.1 Prediction model

We built the HHAN-DSI model for drug-SE prediction using biologically guided propagation and node aggregation (**Fig. 1, Method 4.1**). We first constructed the knowledge graph containing multimodal interactions among drugs, SEs, genes, and gene ontology (GO) (**Fig. 1**, **Method 4.2.1, Table 5**). We then derived meta-paths that customize the trajectory of information propagation in GNN models in pre-defined biological semantics (**Table 1**). We constrained the information propagation among these multimodal interactions by ontological relations or meta-paths (**Table 1**, **Method 4.1.1**). To preserve the biological context of each meta-path, we built separated encoders for each entity and derived node embedding within relevant meta-paths (**Method 4.1.2**), then aggregated the embedding of the underlying entity to the higher level of entities. The first-level hierarchical order was from gene to drug/SEs, and the second-level one was from drugs to SEs, reflecting the fact that an organism’s gene (or drug) interactions can be a basis to learn drug/SE’s (or SE’s) interaction to the organism, but not vice versa. We trained the model so that the node embedding can reconstruct the existing edges and also extrapolate to unknown edges between drugs and SEs. The model’s decoder calculated the TransE^33^ score to make the embedding satisfy arithmetic relations of *entity <drug> + relation<drug associated with SE> ∼ entity<SE>* (**Method 4.1.3**). During optimization, we optimized the TransE score between drugs and SEs in a learning-to-rank framework (**Method 4.1.4**), so that the embedding of drugs and SEs can reconstruct existing drug-SE edges in the knowledge graph and also extrapolate to unknown drug-SE edges.

**Fig. 1:**
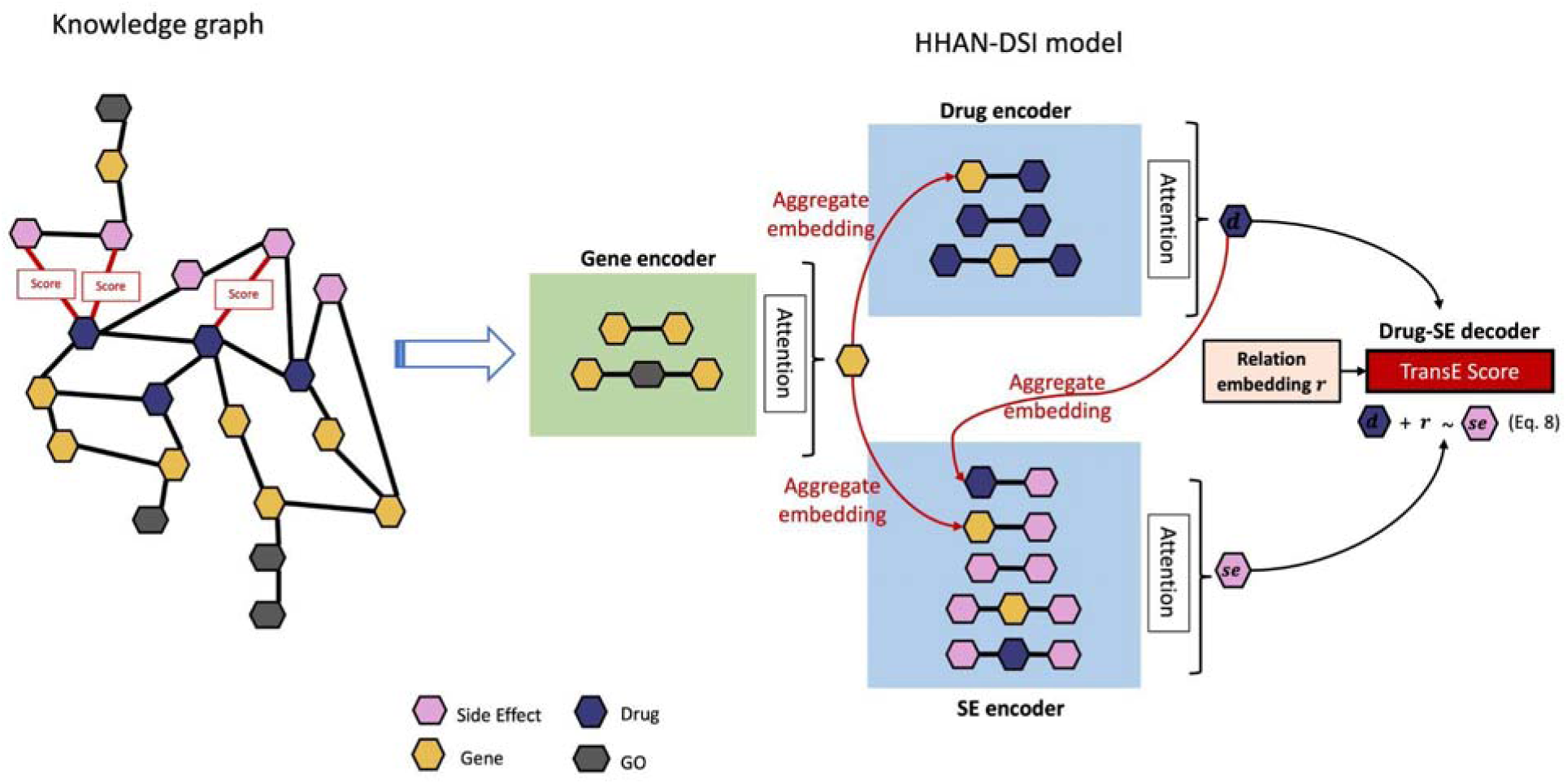
Side effect prediction workflow. The knowledge graph contained multimodal interactions among drugs, SEs, genes, and GOs. HHAN-DSI is a graph neural network architecture that encodes different biological entities into a graph representation with biologically guided propagation and aggregation. Each encoder derives node embeddings within relevant meta-paths. The node embedding flows from genes to drugs and SEs, then from drugs to SEs. The final outcome is a TransE score for each drug-SE pair. Given a drug of interest, SEs with high TransE scores are more likely to occur.

**Table 1:**
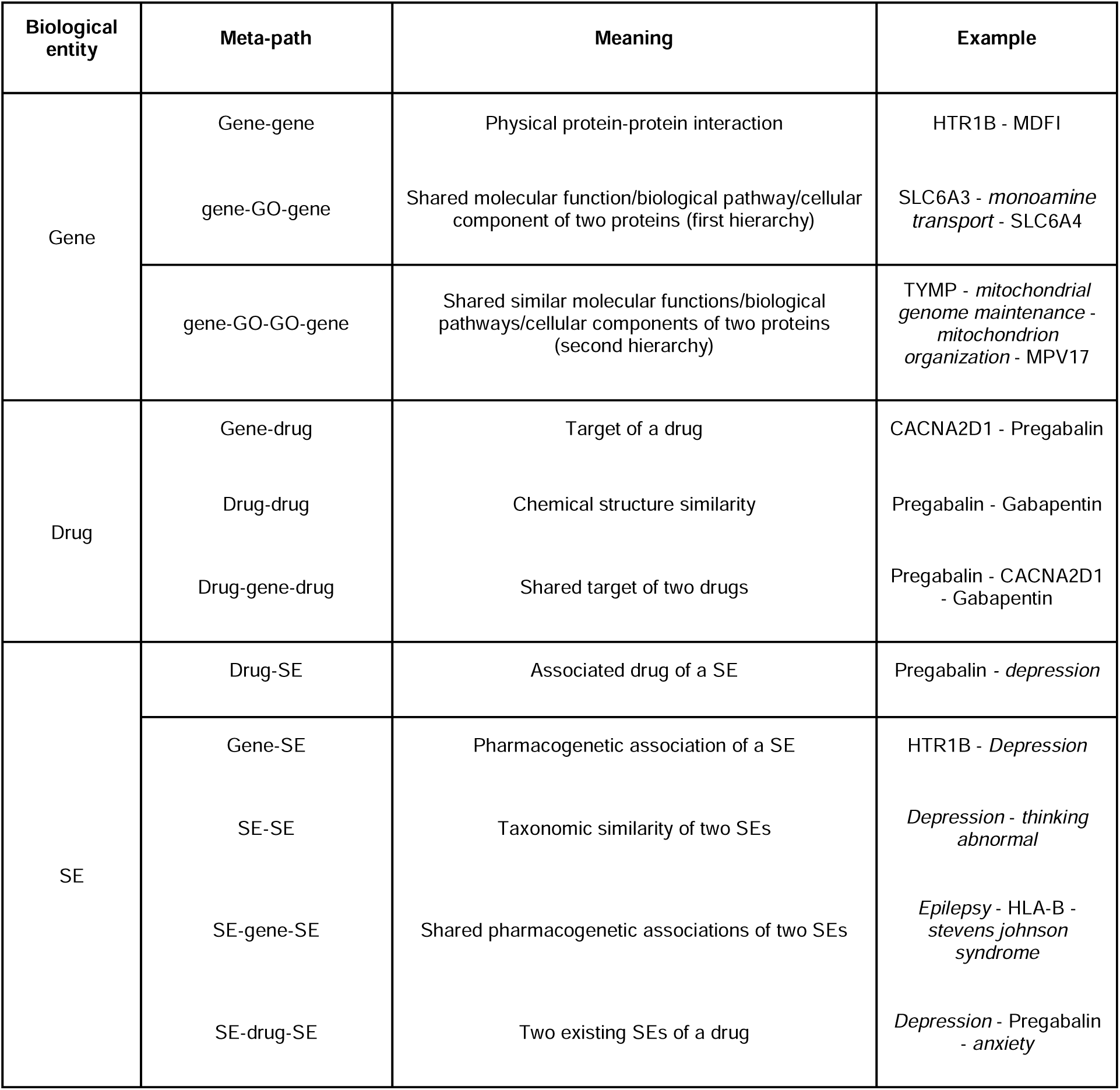
Meta-path types and biological meaning.

#### 2.1.3 Accuracy comparison

We evaluated our model with benchmark data containing a wide range of SEs (Zhang’s data^11^), and compared accuracy with that of the state-of-art works provided by Munoz et al.^34^ (**Table 2**, **Method 4.2.2**). Benchmark experiments had two evaluation scenarios: one with only frequent SEs and one with all SEs.

**Table 2:**
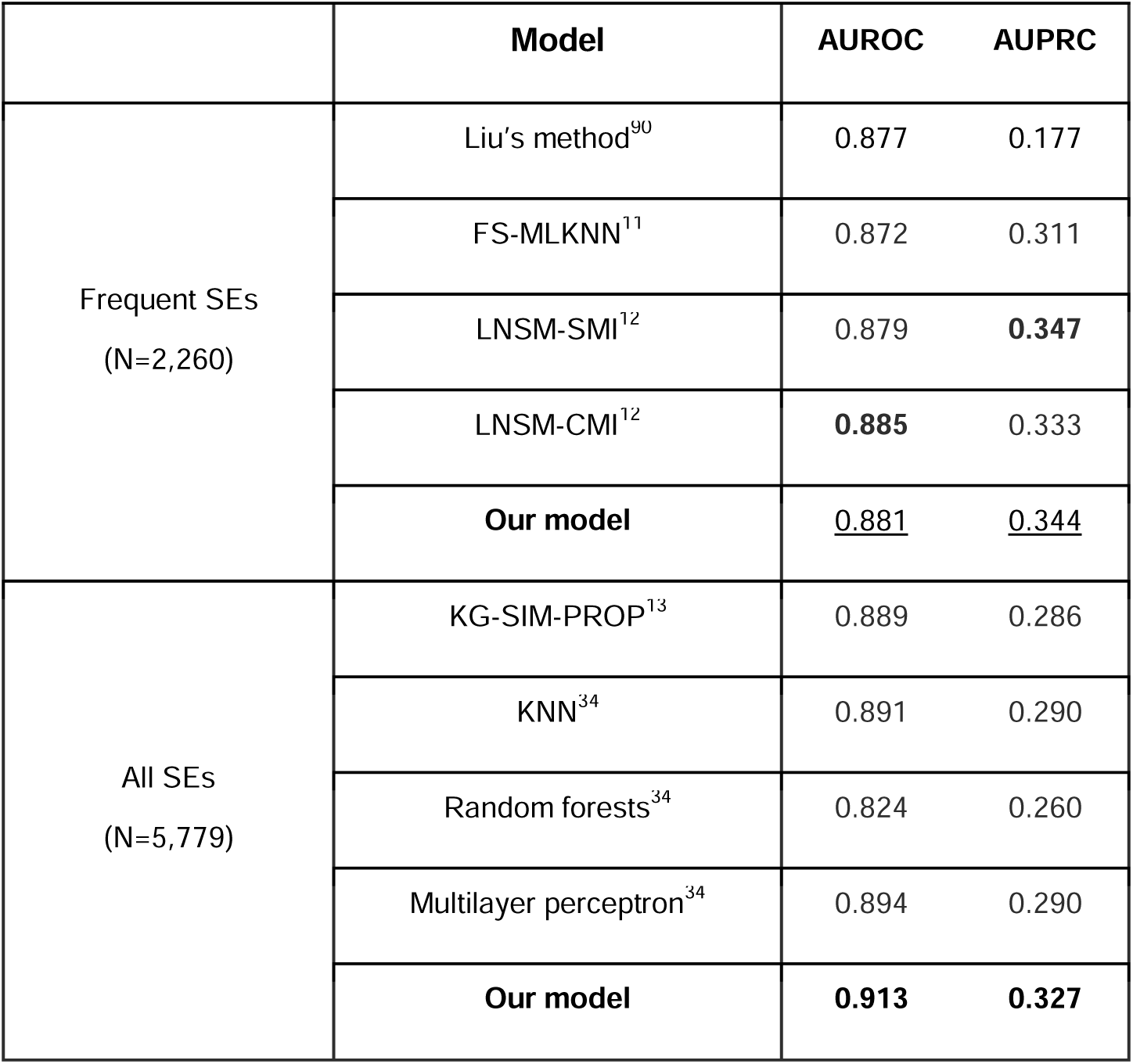
Benchmark comparison of general side effects with Zhang’s data. Each model predicted unseen SEs on the same split of 1,080 drugs. Liu’s method, FS- MLKNN, LNSM-SMI, and LNSM-CMI only predicted 2,260 frequent SEs, whereas other benchmark methods predicted 5,779 all SEs^34^.

|  | Model | AUROC | AUPRC |
| --- | --- | --- | --- |
| Frequent SEs<br>(N=2,260) | Liu’s method<sup>90</sup> | 0.877 | 0.177 |
|  | FS-MLKNN<sup>11</sup> | 0.872 | 0.311 |
|  | LNSM-SMI<sup>12</sup> | 0.879 | **0.347** |
|  | LNSM-CMI<sup>12</sup> | **0.885** | 0.333 |
|  | **Our model** | <u>0.881</u> | <u>0.344</u> |
| All SEs<br>(N=5,779) | KG-SIM-PROP<sup>13</sup> | 0.889 | 0.286 |
|  | KNN<sup>34</sup> | 0.891 | 0.290 |
|  | Random forests<sup>34</sup> | 0.824 | 0.260 |
|  | Multilayer perceptron<sup>34</sup> | 0.894 | 0.290 |
|  | **Our model** | **0.913** | **0.327** |

In the frequent SEs scenario, our model achieved AUPRC of 0.344 and AUROC of 0.881, which was comparable to the most accurate model LNSM-SMI and LNSM-CMI while maintaining explainability thanks to hierarchical information aggregation and knowledge graph structure by design. In the all SEs scenario, our model (AUROC =0.913, AUPRC = 0.327) significantly outperformed other baseline models, such as the knowledge graph method KG-SIM-PROP and traditional machine learning approaches. It suggested that our model can even capture the sparse information around relatively uncommon predicted SEs well.

Furthermore, in order to measure the contribution of biological knowledge, we ablated the biological knowledge constraint and compared the accuracy. We trained a model without the constraint (vanilla HAN) with a varying number of training samples. HHAN-DSI achieved better AUPRC in most sample sizes, and the difference was even distinct with smaller sample sizes (**Fig. 2**). This result verified the utility of biologically informed constraints for detecting unseen SEs even when only limited observations are available (e.g., new drugs).

**Fig. 2:**
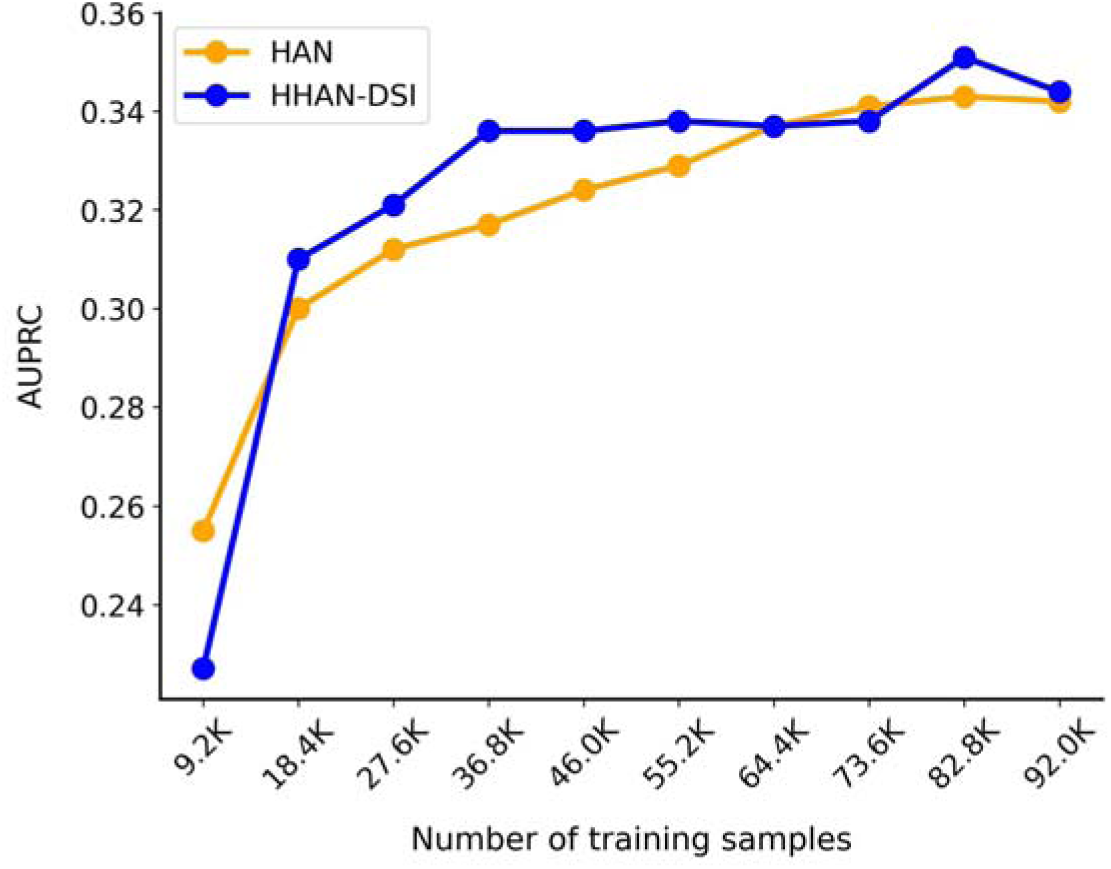
Comparison between HHAN-DSI and one without biologically informed constraint (vanilla HAN) over different sample sizes. We measured the model accuracy of 2,260 frequent SEs based on Zhang’s data.

### 2.2 Case study – predicting CNS-related SEs

After we evaluated our model’s high accuracy, we applied the model to specific types of SEs for in-depth investigation of biological mechanisms. Central neural system (CNS)-related SEs were especially easily overlooked by healthcare providers and can affect a variety of other systems in the body, including musculoskeletal, cardiovascular, respiratory systems, etc., which may result in false psychiatric diagnosis^35^. Therefore, we conducted a case study on predicting CNS-related SEs and explaining the potential mechanisms behind the drug and SE pair.

#### 2.2.1 Data summary

To comprehensively investigate the CNS-related SEs, we extracted them from the latest SIDER^36^ for model training and OFFSIDE^37^ for external validation (Method **4.3.1**). The CNS-related SEs took a large portion of overall SEs. The frequency of CNS-related SEs was skewedly distributed (**Supplementary Fig. 1**). The majority of drug-SE associations were related to the most popular SE, such as *feeling abnormal, paraesthesia, nervous system disorder, confusional state,* and *anxiety*. Less common SEs include *urine odor abnormal*, *leukoplakia oral*, and *vertigo cns origin*. These uncommon SEs can also be very critical and sometimes they cause post-market drug withdrawals^38^. Therefore, we considered predicting all CNS-related SEs, including uncommon ones.

#### 2.2.2 CNS-related side effect prediction results

##### Node Embeddings in UMAP Visualization

We built a SE prediction model focusing on the CNS system. We first investigated whether our model generated node embeddings that preserve multimodal interactions. We visualized output node embeddings using Uniform Manifold Approximation and Projection (UMAP)^39^ to examine the model’s capability in preserving biological context with similar entities. The node embeddings were well grouped by similar biological entities (**Fig. 3**), implying that these node embeddings captured the topology of the knowledge graph. Many nodes in the margin of groups had larger Jaccard index values (**Supplementary Fig. 2**), which means that these nodes are similar to each other. For example, neural-related SEs and blood-related SEs were located at the margin respectively. Azatadine, Chlorphenoxamine, and Clidinium, which were at the margin of the drug node group, were all correlated via drug-drug similarities or target similarities. TACR2, TACR3, TAC3, OXTR, CXCR2, and GRM8 were also closely located at the margin of the gene node group, and they were correlated because they can be affected by the same drugs. In all, the UMAP visualization demonstrated that our model preserved the biological semantics in the node embedding.

**Fig. 3:**
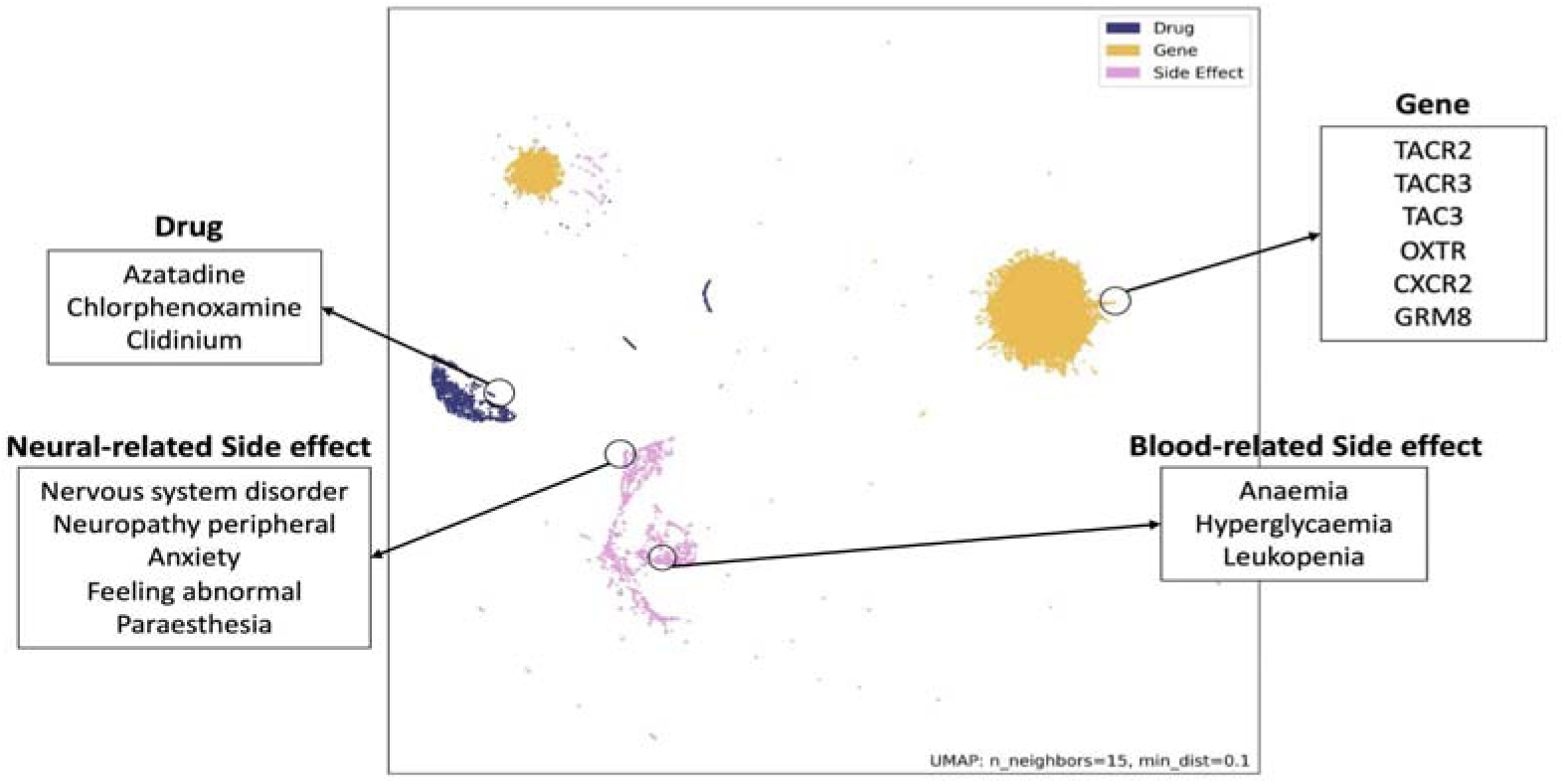
Node embedding via UMAP visualization. Each point shows the position of a node’s 2-dimensional UMAP embedding derived from HHAN-DSI outputs. Different colors show the node type. We annotated some nodes that have high Jaccard Index values, which means that they are similar to each other.

##### Ranking side effects

After checking the validity of node embeddings, we evaluated the accuracy of predicting SEs of drugs by both internal and external validation. We performed internal evaluation to measure the model’s accuracy to predict unseen drugs’ SEs in the same SIDER dataset (**Fig. 4a**). We obtained internal AUROC at 0.945 and AUPRC at 0.364 when training and testing the model with SIDER data, which is higher than the accuracy when predicting all types of SEs (**Table 3**).

**Fig. 4:**
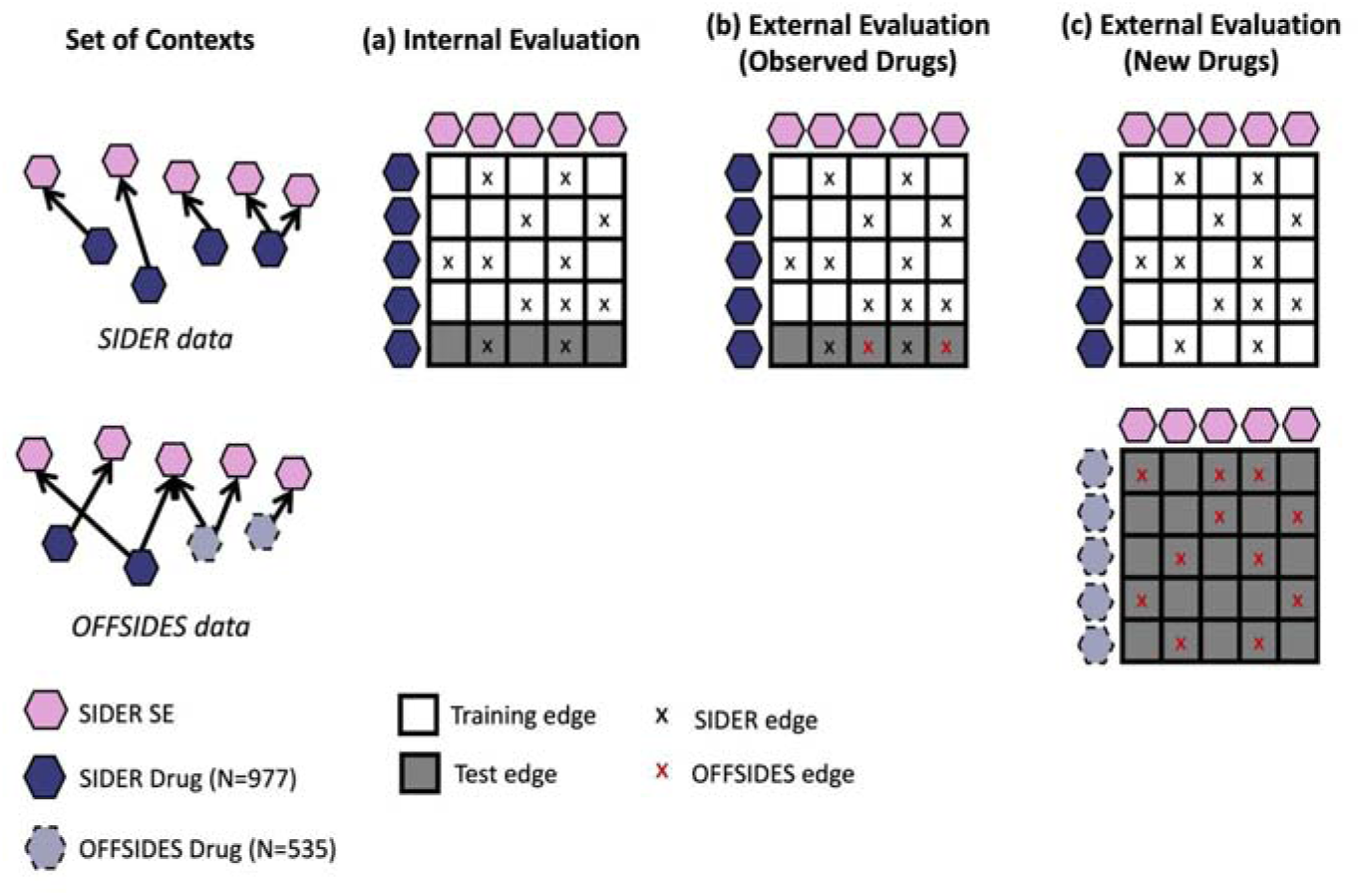
The training and test split of drugs and SEs in SIDER and OFFSIDES. Each row and column stands for a drug and a SE, respectively. Drug-SE pairs used for training our graph model are denoted as white, and pairs used for testing are denoted as gray. The color of the cross labels denotes the original source of the edge.

**Table 3:**
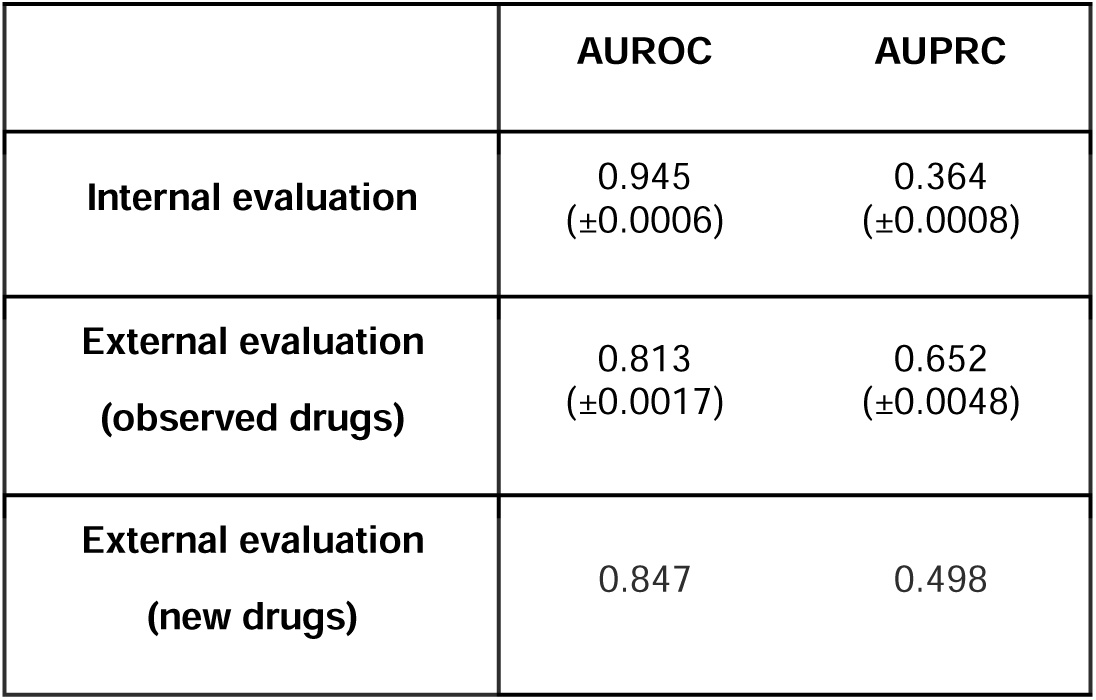
Internal evaluation using SIDER data and external validation using OFFSIDES data.

|  | AUROC | AUPRC |
| --- | --- | --- |
| **Internal evaluation** | 0.945<br>(±0.0006) | 0.364<br>(±0.0008) |
| **External evaluation**<br>**(observed drugs)** | 0.813<br>(±0.0017) | 0.652<br>(±0.0048) |
| **External evaluation**<br>**(new drugs)** | 0.847 | 0.498 |

We performed external evaluation using the OFFSIDES dataset to investigate the accuracy of our model when predicting SEs that were discovered after the drugs were released to the market. This evaluation was intended to emulate a realistic prospective validation as the post-market information is only used in a test set^40^ (**Fig. 4b, 4c**). We externally validated our model under two realistic scenarios: i) to predict a drug’s new SEs when the drug has some known SEs (observed drugs, **Fig. 4b**) and ii) to predict a drug’s SEs when the drug has no known SEs previously (new drugs, **Fig. 4c**). In the first scenario, our model predicted the known drug’s new SEs at AUROC of 0.813 and AUPRC of 0.652. In the second scenario, our model predicted new drugs’ SEs at AUROC of 0.847 and AUPRC at 0.498. The high accuracy with new drugs may be attributed to the fact that our knowledge graph can connect the new drugs to biological knowledge via chemical or genomic similarity and well exploit the underlying biological mechanisms behind them.

We investigated the distribution of predicted TransE scores (**Supplementary Fig. 3**). Many negative edges achieved lower scores and others fell into the score distribution of positive edges. These negative edges with high scores are likely at the top rank.

##### Explaining important meta-paths

We reported the overall contribution of the meta-path to SE predictions (**Fig. 5, Method 4.1.2**). We observed that physical gene-gene interactions primarily contributed to the gene encoder. In the drug encoder, the contribution of the drug’s chemical structure similarity (drug-drug) was close to the genetic similarity (including gene-drug and gene-drug-gene), implying that features of chemical structures and drug targets informed the drug altogether. In the SE encoder, gene-SE played a critical role, implying that the final SE embedding may learn to derive meaningful genomic information from our two-level hierarchical aggregations. SE-drug-SE (i.e., two SEs induced by the same drug) was also important, implying that our model may consider co-occurring SEs as other important candidates when it found a certain SE was highly predictable.

**Fig. 5:**
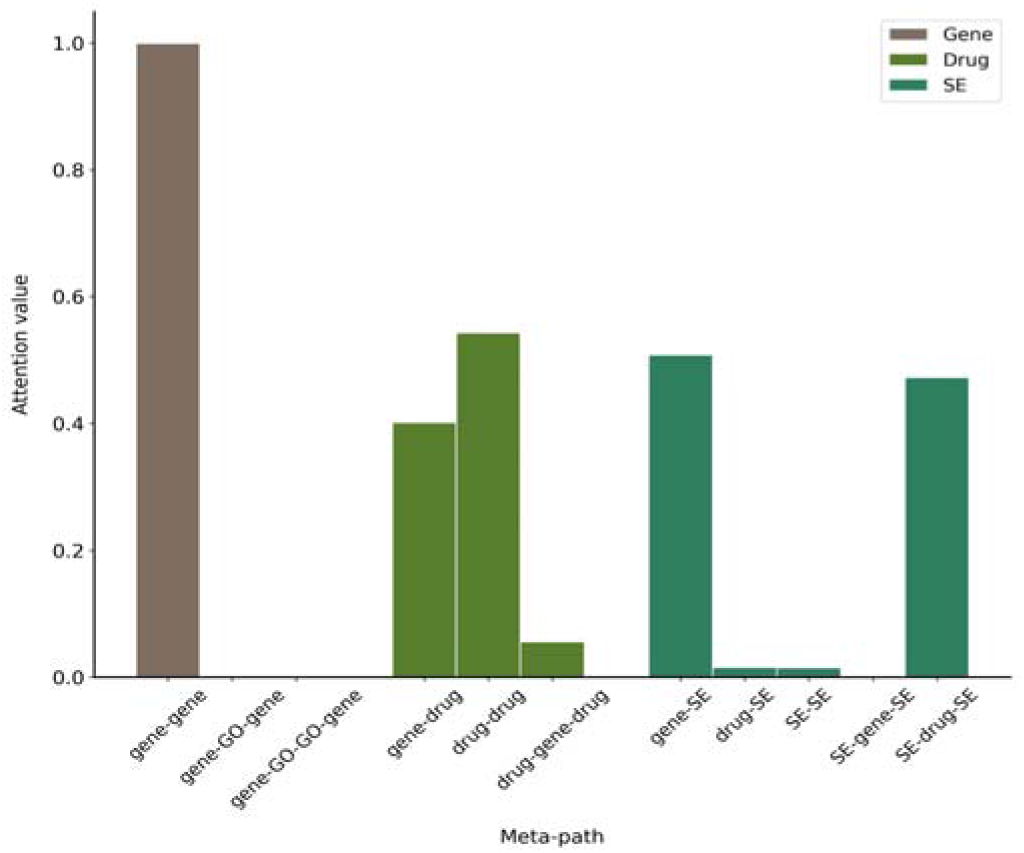
Importance of meta paths when predicting SEs with SIDER data. Since three encoders learned meta-paths within gene/drug/SE, the sum of semantic-level attention values was one in each encoder.

##### Examples of Top-Ranked Drugs and Side Effects

We examined a few drugs that achieved the highest accuracy. We observed that Chlorzoxazone, Praziquantel, Pyrazinamide, Triethylenetetramine, Tolazamide, and Mebeverine were highly predictable (**Supplementary Table 1**), possibly because these drugs had rich pharmacogenomic and chemical information in our knowledge graph.

We evaluated whether drugs of interest and their top-ranked SEs have biological relevance. We focused on Ketamine (AUROC = 0.90) – a drug used medically for induction of anesthesia, pain relief, and antidepressant. However, Ketamine has been reported to induce psychiatric SEs in 10%-20% adults^41^, ranging from *dysphoria* to *hallucinations* and *delirium*^42^, and the long-term effects of regular use are largely unknown. . We observed that many top-ranked SEs have strong and apparent molecular underpinnings with the CNS system. We highlighted the top-10 predicted SEs for Ketamine that were not recorded in package inserts (i.e., SIDER) but reported in post-market surveillance data (i.e., OFFSIDE) with their overall prevalence in OFFSIDES (**Table 4, Supplementary Table 2**). According to the Council for International Organizations of Medical Sciences (CIOMS)’s definition on the frequency of adverse events^43^, most of the top-10 SEs were frequent (1%-10%) and some were infrequent (0.1%-1%). We can see that top-10 predicted SEs were not very common cases (10%-100%) and thus have not been collected in the SIDER database. It suggested that our model has the capability of detecting infrequent and rare SEs.

**Table 4.**
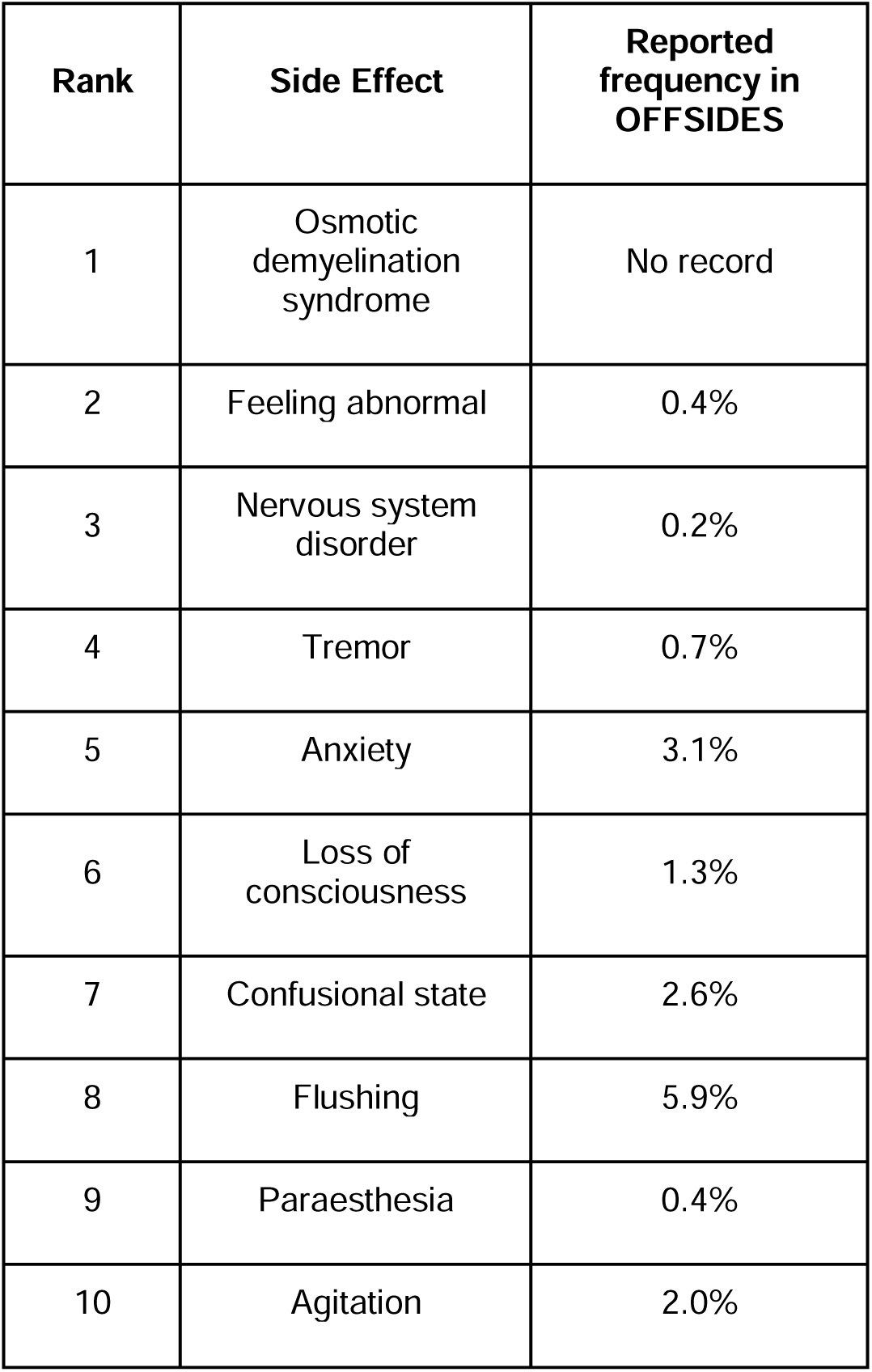
Potential side effects of Ketamine with top ranking scores. Ketamine’s SEs that are not recorded in SIDER but predicted to be likely. They are ranked in descending order. Most new SEs are supported by OFFSIDES records.

#### 2.2.3 Explaining the biological mechanism of SE prediction

To understand the mechanism on how the drug induces SEs, we explained it using the most important interactions and entities that contribute to the prediction using attention weights.

##### Explaining important interactions of drug and side effects

To reveal the contribution of neighbors to the prediction of SEs, we visualized subgraphs (**Fig. 6**; raw images in **Supplementary Fig.1-2**) including important genes, drugs, and SEs that are connected to the nodes of a given drug and SE (**Method 4.1.5**). We chose Ketamine and its top-ranked SEs – *hallucination* (ranked 12nd) and *depression* (ranked 16th) as the visualization example. The subgraph was trained with SIDER data, in which Ketamine was excluded.

**Fig. 6:**
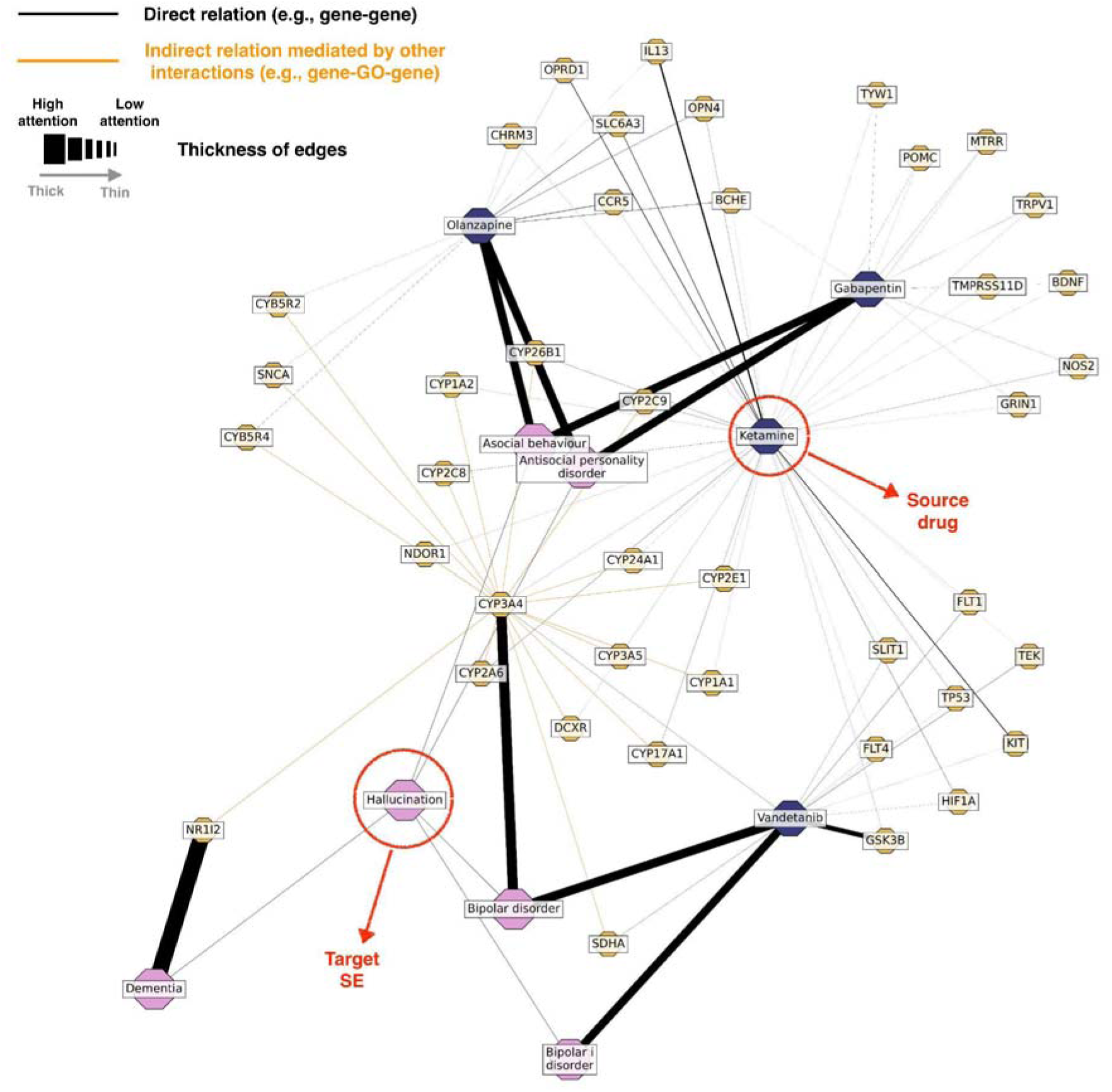
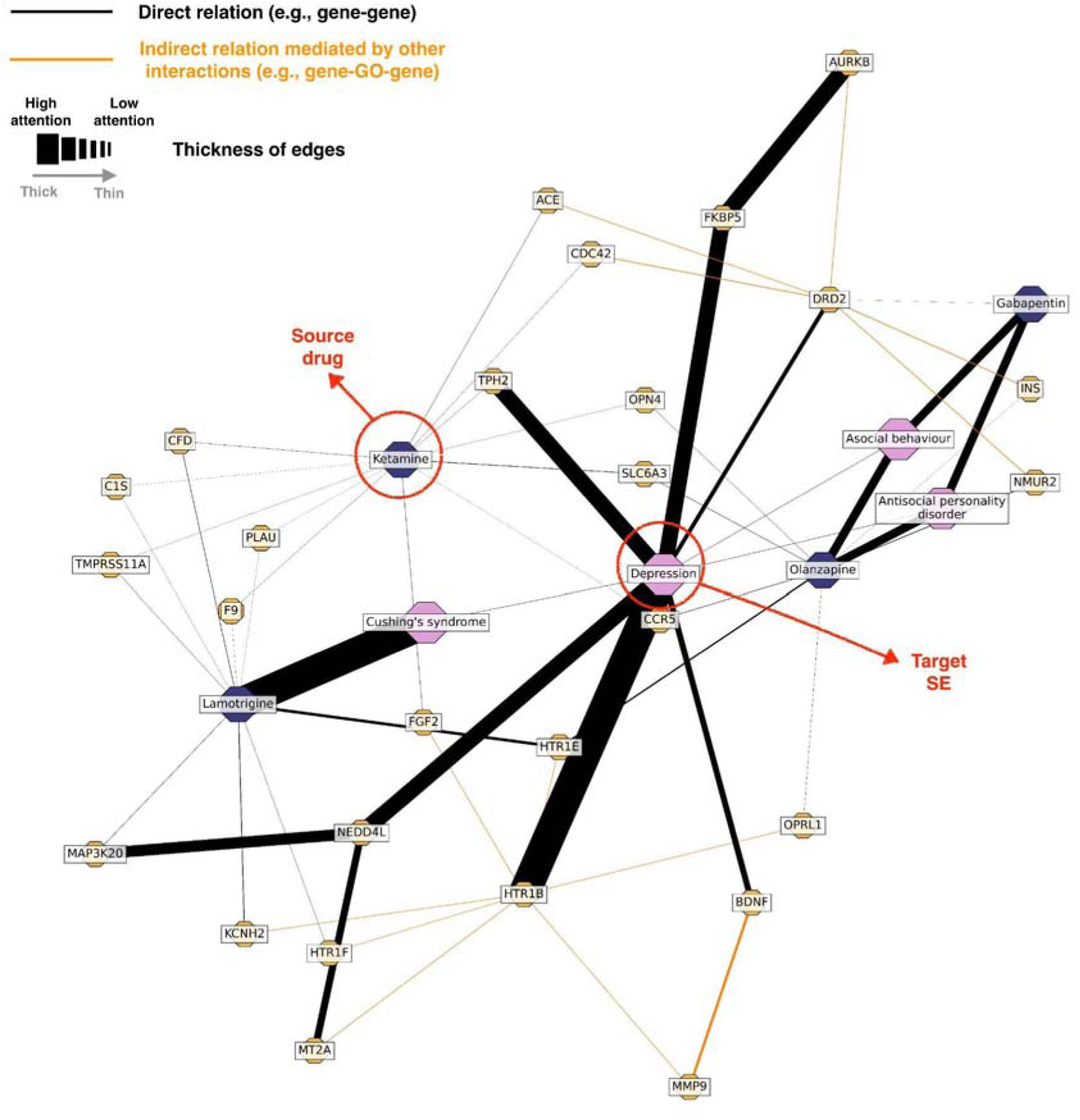
Important subgraphs that explain how Ketamine induces *hallucination* and *depression*, respectively. (a) Subgraph from the *hallucination*’s perspective. (b) Subgraph from the *depression*’s perspective. It explains how neighbors’ pharmacogenomic associations contribute to SE predictions. To limit the graph size, we only presented edges whose weight values were more than the user-defined threshold.

**Fig. 6(a)** represents a subgraph from the perspective of *hallucination*. Ketamine is a so-called ‘dissociative’ anesthetic drug, because it may produce functional and electrophysiological dissociation. To explore the potential mechanism by which Ketamine induces a typical dissociative experience – *hallucination*, we present the subgraph in which the biological pathways were mediated by critical genes or drugs.

- <u>Ketamine-CYP3A4-*bipolar disorder*-*hallucination*:</u> Ketamine can be metabolized into its active form Norketamine by several cytochrome P450 (CYP) enzymes, which increase the drug’s effects on the human body^44^. This reaction is mainly metabolized by CYP3A4, and there is evidence to suggest that CYP3A4 variations may increase the risk of *bipolar disorder*^45^. Severe *bipolar disorder* often leads to *hallucination*, particularly during manic or mixed episodes^46^.
- Ketamine-mechanisms in similar drugs (e.g., Olanzapine/Gabapentin)-*asocial behavior*/*antisocial personality disorder*-*hallucination*: Our subgraph involves similar drugs that share common chemical structures or many target genes. The SE prediction is partly based on the intuition that similar drugs may share similar mechanisms that lead to certain SEs. Our subgraph shows that Olanzapine and Gabapentin are genetically similar drugs that are potentially associated with *hallucination*. There have been some case reports of *hallucination* while taking Olanzapine^47^ or Gabapentin^48^. Both drugs may interfere the individual’s ability to engage in social activities because of changes in the normal activity of neurotransmitter systems in the brain, which may in turn increase the risk of developing *hallucinations*. Olanzapine works by blocking certain receptors in the dopamine (e.g., SLC6A3, **Figure 6(a)**) system, while Ketamine decreases the expression of SLC6A3. Dopamine controls the sense of reality, and too much^49^ or too little^50^ dopamine can cause *delusions* and *hallucinations*. Both Gabapentin and Ketamine may impact the activity of the endogenous opioid system, of which POMC (**Figure 6(a)**) and its associated peptides are a part. Particularly, POMC produces beta-endorphins – the primarily agonist of mu opioid receptors which impacts brain reward-system pathways and social behaviors^51, 52^. While beta-endorphins have not been directly linked to hallucinations, alterations in endogenous opioid signaling have been associated with some psychiatric conditions that may involve *hallucinations*^53^.

**Fig. 6(b)** represents a subgraph from the perspective of *depression*. Ketamine has contradictory influences on *depression*: the low-dose infusion properly administered by the doctor can treat some patients with *depression*, but on the other hand, the long-term use of Ketamine can somehow cause severe *depression*^54^. The subgraph shows how Ketamine causes the adverse effect *depression* with multiple pathway evidence.

- <u>Ketamine-TPH2-*depression*:</u> Ketamine is directly linked to TPH2, which plays an essential role in the physiological processes of serotonin. Specifically, variations in the TPH2 gene have been linked to lower levels of serotonin production, which may contribute to the development of depressive symptoms^55^.
- <u>Ketamine-mediated mechanism (e.g., ACE/CDC42)-DRD2-*depression*:</u> Research has suggested that Ketamine can affect dopamine signaling in the brain, including by blocking N-methyl-D-aspartate (NMDA) receptors, leading to increased dopamine release in the prefrontal cortex. While Ketamine is not a specific DRD2 agonist, its effects on DRD2 are likely indirect^56^. As the major dopamine receptors in the brain, DRD2 is involved in a variety of functions including motor control, cognition, and reward processing. Alterations in DRD2 expression and activity have found in people with *depression*^57^.
- <u>Ketamine-mediated mechanism (e.g., FGF2)-HTR1B-*depression*:</u> HTR1B is one of the several subtypes of serotonin receptors in the brain. Since serotonin receptors are involved in brain functions such as mood regulation and the response to stress, HTR1B has been implicated in the pathophysiology of *depression* and other psychiatric disorders. Ketamine can activate the HTR1B receptor, potentially contributing to its antidepressant effects^58^. On the other hand, however, *depression* is a complex and multifaceted disorder, and certain genetic variations in HTR1B may increase the risk of developing treatment-resistant *depression*^59^.
- <u>Ketamine-mechanisms in similar drugs (e.g., Lamotrigine)-*Cushing’s syndrome*-</u> *<u>depression</u>*: Lamotrigine and Ketamine are genetically similar drugs because they have several overlapping targets. Lamotrigine can lead to *Cushing’s syndrome* due to excessive cortisol production. While our data does not show the interaction Ketamine-*Cushing’s syndrome*, research has shown that Ketamine also has an appreciable effect on cortisol production^60^, potentially contributing to *Cushing’s syndrome*. Elevated cortisol levels can affect the brain and lead to changes in mood and behavior, and many cases show that *depression* is the major psychiatric disturbance in *Cushing’s syndrome*^61^.

## 3. Discussion

We proposed a graph representation model HHAN-DSI to predict SEs of unseen drugs, and to explain the mechanism behind the association between the drug and SE by visualizing important trajectories. Our model processed a multimodal knowledge graph consisting of various biological interactions among drugs, SEs, genes, and GOs. HHAN-DSI propagated node embeddings in guidance of biological nature, allowing for better explainability. HHAN-DSI obtained higher accuracy compared to several baseline models even when SEs are uncommon. Focusing on the CNS system, we emulated a prospective validation by predicting SEs that were discovered after market release.

HHAN-DSI achieved competitive accuracy even with new drugs that are not observed during training. The high accuracy in external evaluations suggests that our model, which is trained with small but high-quality SIDER data (i.e., recorded in package inserts), was well generalized to the sparse and noisy post-market surveillance data in OFFSIDES, thus demonstrating that our model has high utility in the real world. Our model revealed the mechanism behind established SEs (e.g., Ketamine-*hallucination*) and novel SEs (e.g., Ketamine-*depression*). The explainable subgraph suggested that the drug may induce SEs due to their similar pharmacogenomic activities.

HHAN-DSI was designed to encapsulate multimodal and complex molecular interactions in a biologically meaningful way. We aggregated information from the lower level of molecular mechanisms (interactions of genes) to the higher level (interactions of drugs and SEs), and restricted multi-hop neighbors to derive biologically meaningful pathways from existing one-hop neighbors. Thanks to biological constraints, our model was robust on sparse data, including the limited sample size and uncommon SEs. In addition, our model’s strong capability of detecting uncommon SEs is also because of several relation types between SEs (SE-drug-SE, SE-SE) such that uncommon SEs can share common parameters with popular SEs.

Besides, HHAN-DSI allows for local and global biological interpretation across multiple interaction modalities. Globally, we found that most direct relation types (gene-gene, gene-drug, drug-drug, gene-SE) and some mediated indirect relation types (drug-gene-drug, SE-drug-SE) contributed the most in predicting SEs (**Fig. 5**). These patterns recapitulate the current knowledge of chemical functional groups (drug-drug)^12, 62^, common protein targets (drug-gene)^12, 63^, and pharmacogenetic associations (gene- SE)^64, 65^, as well as the protein interaction network (gene-gene)^66^, which can importantly impact whether a drug causes a certain SE. Few studies also validated the contribution of co-occurring SEs (SE-drug-SE), thus co-occurring SEs may be a novel but critical feature. Our results suggested that some co-occurring SEs were potentially correlated because of their function sites on the common tissue. Locally, HHAN-DSI can exhibit decisive relation types for predicting both novel and existing SEs of a drug, providing insights on molecular mechanisms or influences of chemical structural relevance on a SE.

However, our work has the following limitations. First, we only leveraged a few gene-SE associations due to the small data size. In our knowledge graph, SE nodes were mainly characterized by categorical and co-occurring SE neighbors. We believe that incorporating more pharmacogenomic information involved in SEs may uncover hidden mechanisms between the drug-SE pair. Second, our knowledge graph takes up a lot of storage space. Our future work will focus on finding high-quality interactions among our data, in order to reduce unnecessary storage and computations. Finally, our prediction model is not personalized based on individual patient’s pharmacogenomic profiles and prescription details, because access to such individual’s SEs records were very limited. Also, distinguishing the mechanisms between drug SEs and indications was difficult in our study, because the molecular action may vary under distinct cohorts or prescription conditions (e.g., underdose, overdose, or unstable status). For example, Nortriptyline is an antidepressant whereas *depression* is reported to be a very common SE. Future studies will incorporate the dosage of medications and the longitudinal development (e.g., manic switch with antidepressant treatments) of SEs.

## 4 Method

### 4.1 Graph Representation Model

The knowledge graph *G* consists of a set *V* of nodes and a set *E* of edges. Node and edges have heterogeneous types with a node type mapping function *ϕ* : *V ➔ A* and an edge type mapping function, *φ: E ➔ R*, where *A* and *R* denote entity and edge types. Our graph *c* includes four entity types (drug, SE, gene, and GO) and a series of relations among them.

We built a graph representation model HHAN-DSI that learns embedding from the knowledge graph. It consists of three heterogeneous attention network (HAN) encoders and a TransE decoder (**Fig. 1**). The HAN encoder derives embedding from the knowledge graph following predefined meta-paths and the hierarchy among entities. The decoder calculated the relevance score (i.e., TransE) among entities and the relation (e.g., a drug causes a SE) using the encoded node embedding. The model was trained to reconstruct the drug-SE edges in the knowledge graph and also inferred unknown drug-SE edges.

#### 4.1.1 Meta-path selection

The meta-path^67^ is widely used to capture the semantics of a knowledge graph. It is in the form of *A*_l_ *➔ A*_2_ *➔…➔ A*_l+l_, which describes the composite relation *R = R*_l_*o R*_2_*o…o R_i_* between different node types *A*_l_ and *A*_l+l_, where the number of subset relations *i* >= 1. The nodes *A*_l_ and *A*_l_ are meta-path neighbors. The original edge type between two nodes is the simplest meta-path. For example, a typical meta-path between two drugs can be defined as *drug ➔ gene ➔ drug*, which means that two different drugs are connected because they target the same gene. *sE ➔ drug ➔ sE* describes that two SEs are connected because they could be caused by the same drug.

#### 4.1.2 Meta-path guided encoder

We built separate encoders for genes, drugs, and SEs, and then learned node embedding within their own meta-paths. First, each type of node was transformed into a unique feature space. Given a knowledge graph G = (*V*,*E*) and the type-specific transformation matrix *M_ϕ_*_i_, the projection process can be shown as follows:

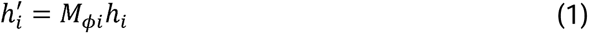

where *h*_i_ and *h_i_*’ are the initial and projected feature of node *i*. Next, HAN uses the node pair (*i*,*j*) in a meta-path *φ*, the importance of a meta-path node pair *c;, 11* can be node-level attention mechanism to learn the weight of meta-path neighbors. Given a formulated as:

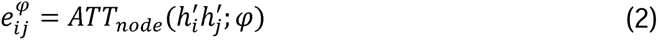

where *ATT*_node_ denotes the node-level attention function, which is shared for all node pairs (*i,j*) within a meta-path *φ*. It is because there are similar biological patterns under one meta-path. After obtaining the importance between node pairs in every meta-path, the normalized attention weight is calculated as:

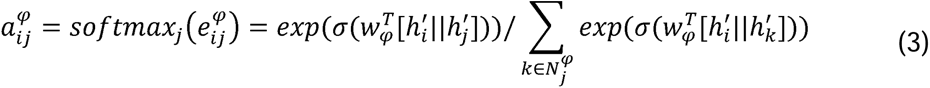

where *σ*, denotes the activation function, *||* denotes the concatenation operator, and *w_φ_^T^* features into node ; with the corresponding multi-head attention weight *α_i,j_^φ^* as follows: is a node-level attention vector. Then, the model aggregates the neighbor’s projected

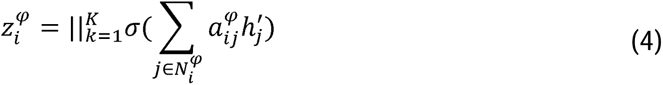

where z*_i_^φ^*is the learned embedding of node *i*, and *K* is the number of attention heads.

After achieving node-level attentions, we learned the semantic-level attention that could be reflected by the meta-path. Taking *P* types of meta-paths {*φ*_1_, *φ*_2_,…, *φ_P_*}, we obtained the importance of each meta-path type *u*_({JP_ as follows:

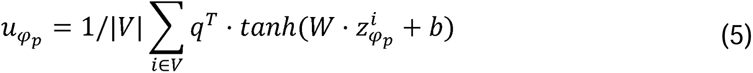

where |*V*| is the number of nodes within the meta-path type, *W* is the weight matrix, *b* is the bias vector *q*, is the learnable semantic-level attention vector. The normalized weight of each meta-path *β_φp_* is formulated as follows:

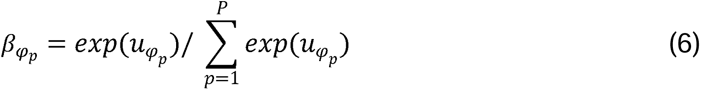

This equation shows the contribution of the meta-path type for SE predictions. Obviously, the higher *β_φp_* means the more important such a meta-path *φ_p_* is. With the learned node-level and semantic-level weights, we can fuse these embeddings to obtain the final embedding *Z*:

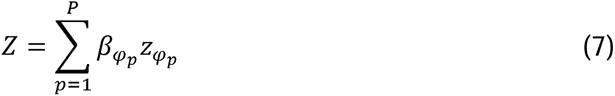

where z*_φp_* is the learned node-level embedding in a semantic group *φ_p_*.

#### 4.1.3 Decoder

Our decoder calculates TransE score^68^, a widely used scoring function in the knowledge graph embedding to measure the relevance of entity and relation. In the embedding space, when adding the relation embedding *r* to the source node *d* (drug entity), we should get close to the target node *se* (SE entity). We used the L1 norm to get the sum of absolute embedding values and added a negative sign to the final value.

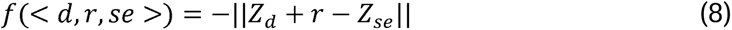

where the relation embedding *r* was randomly initialized in values of [-1, 1].

#### 4.1.4 Learning to rank framework and ranking loss

We formulated the SE prediction as a learning to rank problem, where the model is trained to generate ranking scores (i.e., TransE) of relevant or irrelevant entities (i.e., SEs) given a query entity (i.e., a drug of interest). Positive or relevant entities (i.e., known SEs for the query drug) should get higher ranking scores than negative or irrelevant entities (i.e., random SEs for the query drug). **That is, the drug of interest is likely to cause the SE if its ranking score is high.**

We adopted pairwise hinge loss of TransE scores^69^ to distinguish positive edges from negative edges as an objective function to minimize:

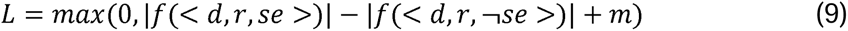

where *f*(*< d,r,se >*) denotes the TransE score of the positive edge, f(< *d*, *r*, ┐*se* >) denotes the TransE score of the negative edge, and *m* represents the customized margin value which controls the marginal gap between score distributions of positive and negative samples. This loss function maximizes differences between the score of positive and negative edges by a reasonable margin *m*.

#### 4.1.5 Generating explainable subgraphs

Once getting the optimized knowledge graph representation, we investigated important interactions involved in predicting potential SEs of a drug of interest. We extracted attention weights learned by HHAN-DSI to describe the importance of neighbor nodes that are aggregated to the target node (**Equation 3**). The higher attention value indicates the greater contribution to a target node, and might significantly influence the final SE prediction. The subgraph consists of several fully-connected paths that depart from the query drug, and recursively connected important neighbor nodes until reaching the query SE. Since the visualized subgraph was undirected, all types of relations and meta-paths could be part of the subgraph. To obtain a compact view, the subgraph did not contain leave nodes other than the query drug and SE.

### 4.2 Benchmark comparison

To measure the model’s generalizability, we used Zhang’s data^11^ to predict a variety of general SEs.

#### 4.2.1 Data source

Zhang’s data^11^ collected 1,080 drugs and 5,779 SEs, and integrated six types of features covering the enzyme, pathway, target, transporter, treatment, and substructure. The benchmark methods used original labels and six features in Zhang’s data. While implementing our model, we built our own features (**Fig. 1**) to predict original labels on Zhang’s data.

Our knowledge graph involves multimodal interactions among biological entities.

Biological interactions in the knowledge graph are described in the following.

##### Gene-related interactions

We focused on experimentally validated physical interactions in human, excluding genetic and indirect interactions between genes. We mapped genes to human biological functions (34,777 GO-gene edges) they affect by using the human version of the Gene Ontology (GO)^70, 71^. We only allowed experimentally validated gene- GO associations according to those referred from experiments, assays, phenotypes, and genetic interactions^72^. We constructed a hierarchy of biological functions (22,545 GO-GO edges) by using Gene Ontology’s Biological Process^71, 73^. The hierarchical relationship between biological functions was curated, where specific biological functions are children of more general biological functions.

##### Drug-related interactions

All drugs were linked to DrugBank^74^. We mapped drugs to their protein targets (334,631 gene-drug edges) using Search Tool for Interactions of Chemicals (STITCH)^75^, DrugBank^76^, and Drug Repurposing Hub^77^. Drug-drug structural similarities (35,922 drug-drug edges) were identified by RDKit^78^, which calculates the similarity of given two compounds using four types of molecular fingerprints, including atom-pair fingerprints, Molecular ACCess System (MACCS) fingerprint, Morgan/Circular fingerprint, and topological-torsion fingerprints. For each fingerprint type, we calculated the Sorensen– Dice coefficients and normalized them to z-scores, then set a threshold of z-score >= 3 for drug-drug similarity associations. A network of physical interactions (387,626 gene-gene edges) was integrated from several major databases^79–86^.

##### SE-related interactions

We mapped drug-induced SEs to genetic variants (880 gene-SE edges) using clinical annotations in PharmGKB^87^. All gene-SE associations were curated according to multiple literature evidence. We constructed the SE-SE taxonomic similarity (98,338 SE- SE edges) by using the MedDRA terminology classification^88, 89^. The relationship between SE terms is present if they belong to the same Main or HLT MedDRA category.

#### 4.2.2 Baseline models to compare

For comparison, we used the following state-of-art models for SE prediction as baselines:

**Liu’s method**^90^: The support vector machine (SVM) classifier for each SE. It integrated drug features from multiple sources, including chemical (substructure), biological (target, transporter, enzyme, pathway), and phenotypic (treatment indication, other SEs) information.
**FS-MLKNN**^11^: The feature selection-based multilabel k-nearest neighbor. This method found critical feature dimensions using the mutual information, and then selected a subset of features to construct an ensemble of five MLKNN models using the genetic algorithm.
**LNSM-SMI**^12^: The linear neighborhood similarity method using similarity matrix integration. It assumed that a drug could be optimally constructed by using a linear combination of its neighboring drugs. This method generated K similarity matrices from K feature types, and integrated similarity matrices using learning weights.
**LNSM-CMI**^12^: The linear neighborhood similarity method using cost minimization integration. The assumption of drug construction was the same as LNSM-SMI. This method learned the similarity matrix independently from each feature type.

The optimal feature type was that had minimum cost during the training stage.

- **KG-SIM-PROP :** The knowledge graph similarity propagation. This method built a multi-relation knowledge graph for drugs and other biological entities. It constructed drug clusters using the KNN algorithm, and aggregated SEs from the drug to its close neighbors.

#### 4.2.3 Evaluation criterion

We calculated the area under the receiver-operating curve (AUROC) when predicting TransE scores of present/absent drug-SE edges. Since there were many more negative labels than positive labels, we also analyzed the ranking performance using the area under the precision-recall curve (AUPRC). AUROC and AUPRC both measure the ranking performance by using a list of thresholds to classify predicted TransE scores.

#### 4.2.4 model training

We followed the experiment settings provided by Munoz et al.^34^, where the frequent SE scenario used 2,260 frequent SEs that share more than 3 common drugs, and the all SE scenario used all 5,779 SEs in the dataset. Among 1080 drugs in Zhang’s data, we trained 771 drugs obtained from Liu’s data^90^, and tested 309 drugs extracted from an independent SIDER 4 dataset.

Due to the large size of our graph, we trained and tested the graph model on a smaller subgraph using the neighbor sampling method^91^. The sampled subgraph involved all drug-SE interactions along with numerous neighbors. To generate an informative knowledge graph, we iteratively sampled 2048 neighbors for each node involved in 3 iterations.

We randomly initialized 200-dimensional entity embeddings in values of [-1,1], and derived 200-dimensional output embeddings by updating the message passing and node aggregation behind HHAN-DSI. Then we used the TransE decoder to translate a triplet of drug embedding, relation embedding, and SE embedding into a score. We applied fixed hyperparameters in all training scenarios. For our model, the number of HAN layers in the subgraph encoder was 1, and the dimension of the hidden linear layer was 400. We used an Adam optimization algorithm to train the model for 2,000 epochs. The learning rate was set to be 10e-5.

### 4.3 Case study experiments

To measure the model’s specificability, we used our data (**Table 5**) to predict CNS-related SEs.

**Table 5:** Details about Zhang’s data and our data.

|  | # drug | # Side effect | # gene | # gene ontology |
| --- | --- | --- | --- | --- |
| Zhang's data | 1,080 | 5,779 | 1,050 | N/A |
| Our data | 1,782 | 2,657 | 19,392 | 9,798 |

#### 4.3.1 data source

We mapped CNS-related SE terms on the preferred term (PT) level according to MedDRA dictionaries^92^. We obtained SEs from two adverse event databases: SIDER^93^ containing 2,657 SEs that were curated from public documents and package inserts, and OFFSIDES^94^ containing SEs in common that were obtained from post-market surveillance data in FAERS^95^. OFFSIDES data are likely to be less reliable because they were statistically inferred^96^. Therefore, we built the knowledge graph using SIDER data only, and set aside OFFSIDES for independent validation of SE predictions. In total, our data collected 1,782 drugs, among which 977 drugs have SEs recorded in both SIDER (44,722 drug-SE edges) and OFFSIDES (684,518 drug-SE edges after removing SIDER duplicates), 535 drugs have SEs recorded in only OFFSIDES (183,750 drug-SE edges), and the remaining 270 drugs are used for constructing drug-drug edges.

We used the same multimodal interactions as benchmark experiments (**Method 4.2.1**).

#### 4.3.2 model training

We used the same evaluation criteria as benchmark experiments (**Method 4.2.3**). We trained and explained our graph model on SIDER, and conducted independent external validations on OFFSIDES to measure model generalizability to unseen drug-SE interactions. We illustrated data split details in **Fig. 4**. Both internal evaluation and external evaluation (observed drugs) performed 5-fold cross-validation to split 977 SIDER drugs on the same seed. Internal evaluation trained and tested with observed SEs in SIDER, while external evaluation (observed drugs) trained with observed SEs in SIDER and tested with observed SEs in SIDER and OFFSIDES. Some testing negative edges were changed to positive in external evaluation (observed drugs) because they are only present in OFFSIDES. External evaluation (new drugs) trained the model with 977 SIDER drugs and tested that with 535 OFFSIDES drugs. We reported mean and standard deviation of accuracy on three random split seeds except for external evaluation (new drugs).

## Data Availability

Data availability are as follows: Zhang's side effects (https://doi.org/10.1186/s12859-015-0774-y), package-insert side effects (SIDER 4.1, http://sideeffects.embl.de/), post-marketing side effects (OFFSIDES, https://tatonettilab.org/offsides/), human protein-protein interaction network that contails physical interactions (BioSNAP, https://snap.stanford.edu/biodata/datasets/10000/10000-PP-Pathways.html), Gene Ontology's Biological Process97 (https://github.com/snap-stanford/multiscale-interactome), drug target data (DrugBank, https://go.drugbank.com/), gene-SE interactions (PharmGKB variant annotations, https://www.pharmgkb.org/downloads), SE-SE categorial similarities98 (Main or High-Level Term Medical Dictionary for MedDRA terminology classification for each side effect term). The curated CNS-related dataset is publicly available at https://github.com/th8930/HHAN-DSI.

## Acknowledgments

XJ is CPRIT Scholar in Cancer Research (RR180012), and he was supported in part by Christopher Sarofim Family Professorship, UT Stars award, UTHealth startup, the National Institute of Health (NIH) under award number R01AG066749 and U01TR002062. YK is partly supported by R01AG066749 and RR180012.

## Author Contributions

TH wrote draft and conducted experiments; TH created plots and visualized the finding; KL curated data from publicly available sources and conducted biological validations; KL, RV, JS discussed the biological implication of findings; KL, YK, XJ contributed to the generation of the idea and contributed to the writing of the paper. All authors made multiple rounds of edits and carefully reviewed the manuscript.

## Data Availability

Data availability are as follows: Zhang’s side effects (https://doi.org/10.1186/s12859-015-0774-y), package-insert side effects (SIDER 4.1, http://sideeffects.embl.de/), post-marketing side effects (OFFSIDES, https://tatonettilab.org/offsides/), human protein-protein interaction network that contails physical interactions (BioSNAP, https://snap.stanford.edu/biodata/datasets/10000/10000-PP-Pathways.html), Gene Ontology’s Biological Process^97^ (https://github.com/snap-stanford/multiscale-interactome), drug target data (DrugBank, https://go.drugbank.com/), gene-SE interactions (PharmGKB variant annotations, https://www.pharmgkb.org/downloads), SE-SE categorial similarities^98^ (Main or High-Level Term Medical Dictionary for MedDRA terminology classification for each side effect term). The curated CNS-related dataset is publicly available at https://github.com/th8930/HHAN-DSI.

## Code Availability

Python implementation of our methodology is available at https://github.com/th8930/HHAN-DSI. All analyses were performed using Python 3.7, Pytorch 1.12, PyTorch Geometric 2.0.4, NetworkX 2.3. Please read the README file at the GitHub repository for information on additional packages and how to run the code.

