## Supplementary Material for "Explainable drug side effect prediction via biologically informed graph neural network"

Supplementary Information includes:

Supplementary Figures **1**-**3** in this document

Supplementary Tables **1**-**2** in additional excel files

- **S Table 1.** AUROC of each drug of interest under internal evaluation scenario
- **S Table 2.** Potential side effects of Pregabalin with ranking scores and ground truth SIDER/OFFSIDES labels (1 = present; 0 = absent)

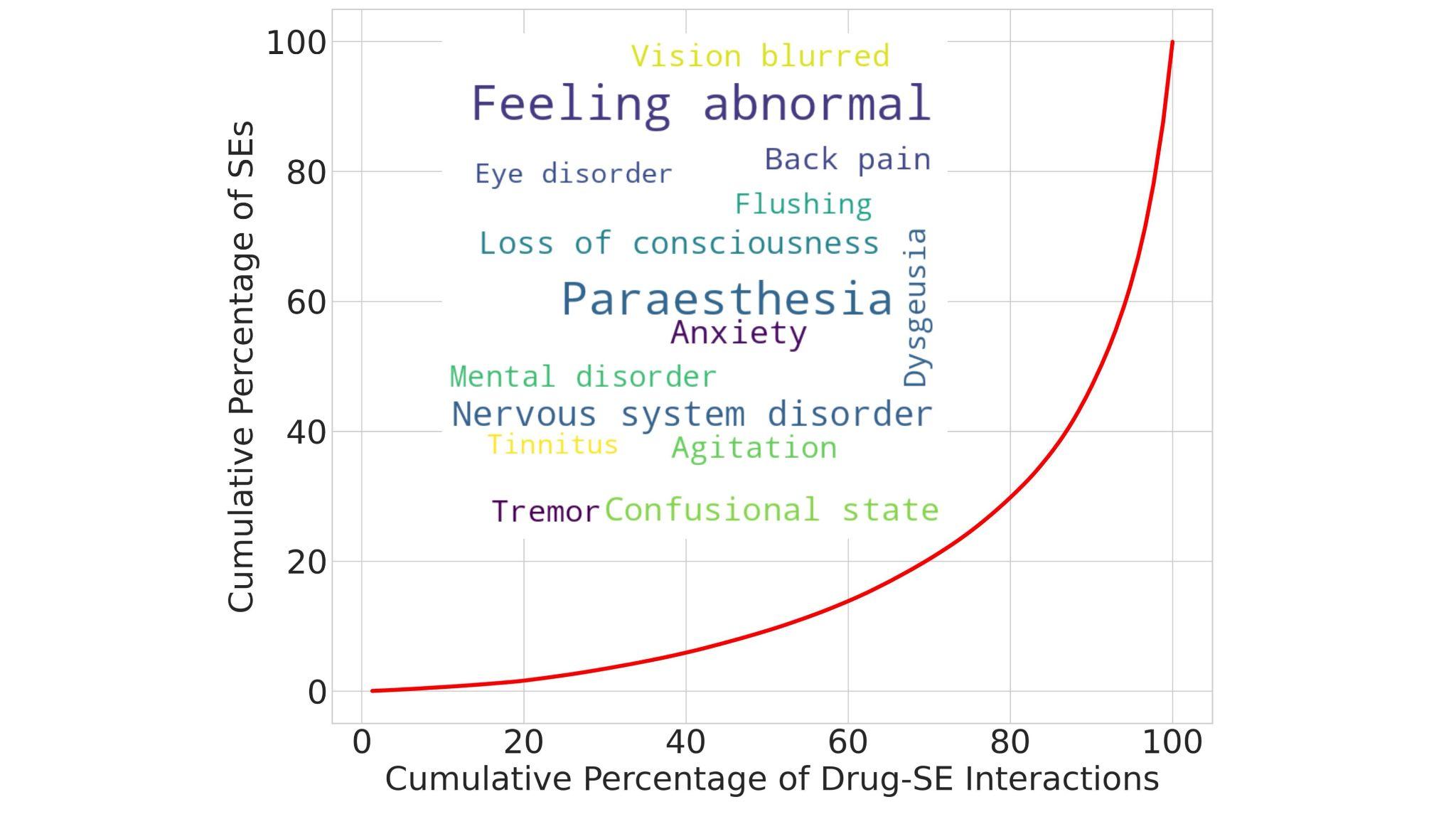

**Supplementary Fig. 1: Distribution of SE terms.** The word cloud in each histogram presents the top-15 most popular terms. The word size is proportional to its frequency.

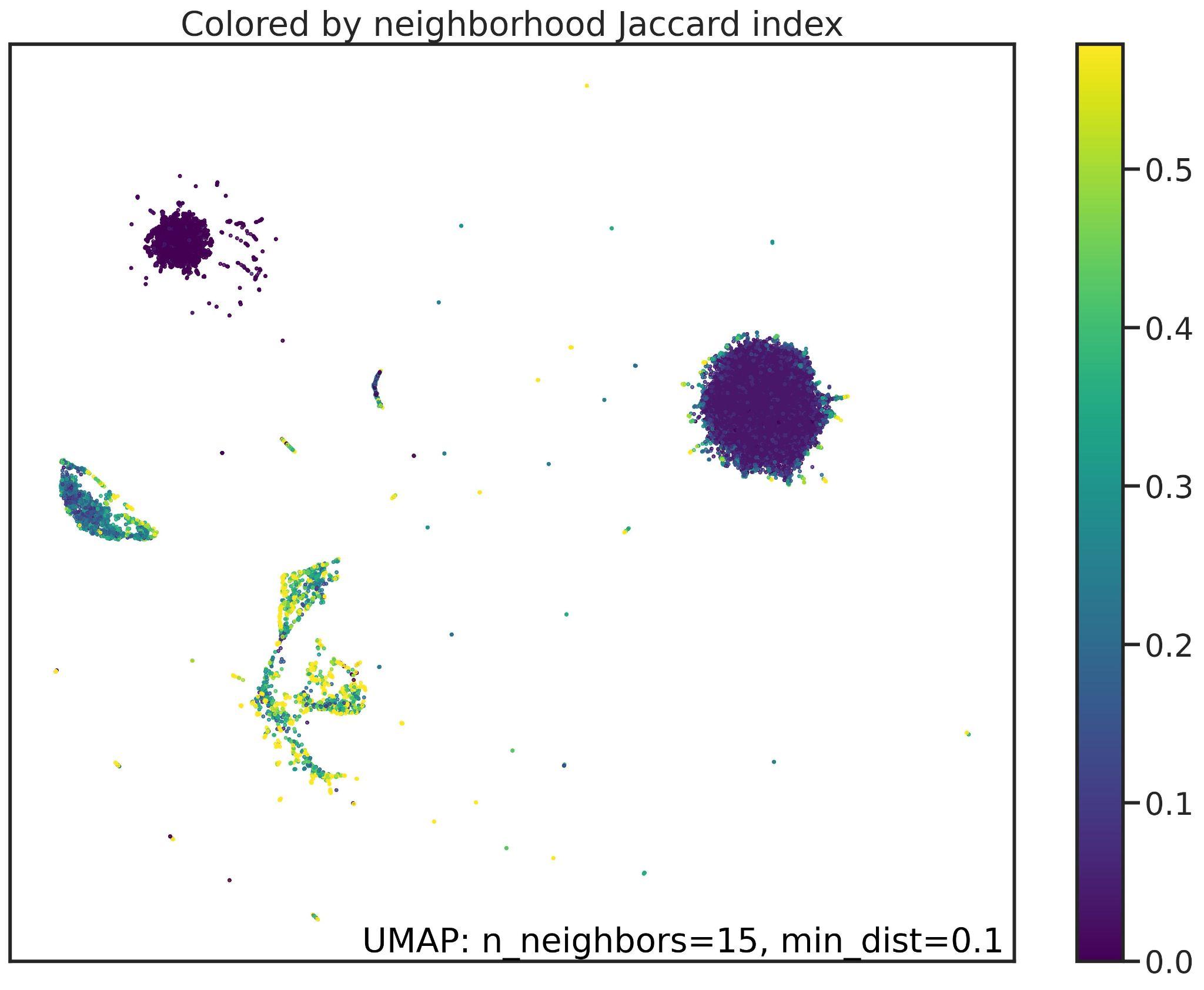

**Supplementary Fig. 2: Jaccard index plot on node embeddings.** Jaccard index is the proportion of the number of neighbors that the two nodes have in common over the total number of distinct neighbors across the two nodes. The Jaccard index is high if two nodes share many neighbors.

| (a) **Internal Evaluation**   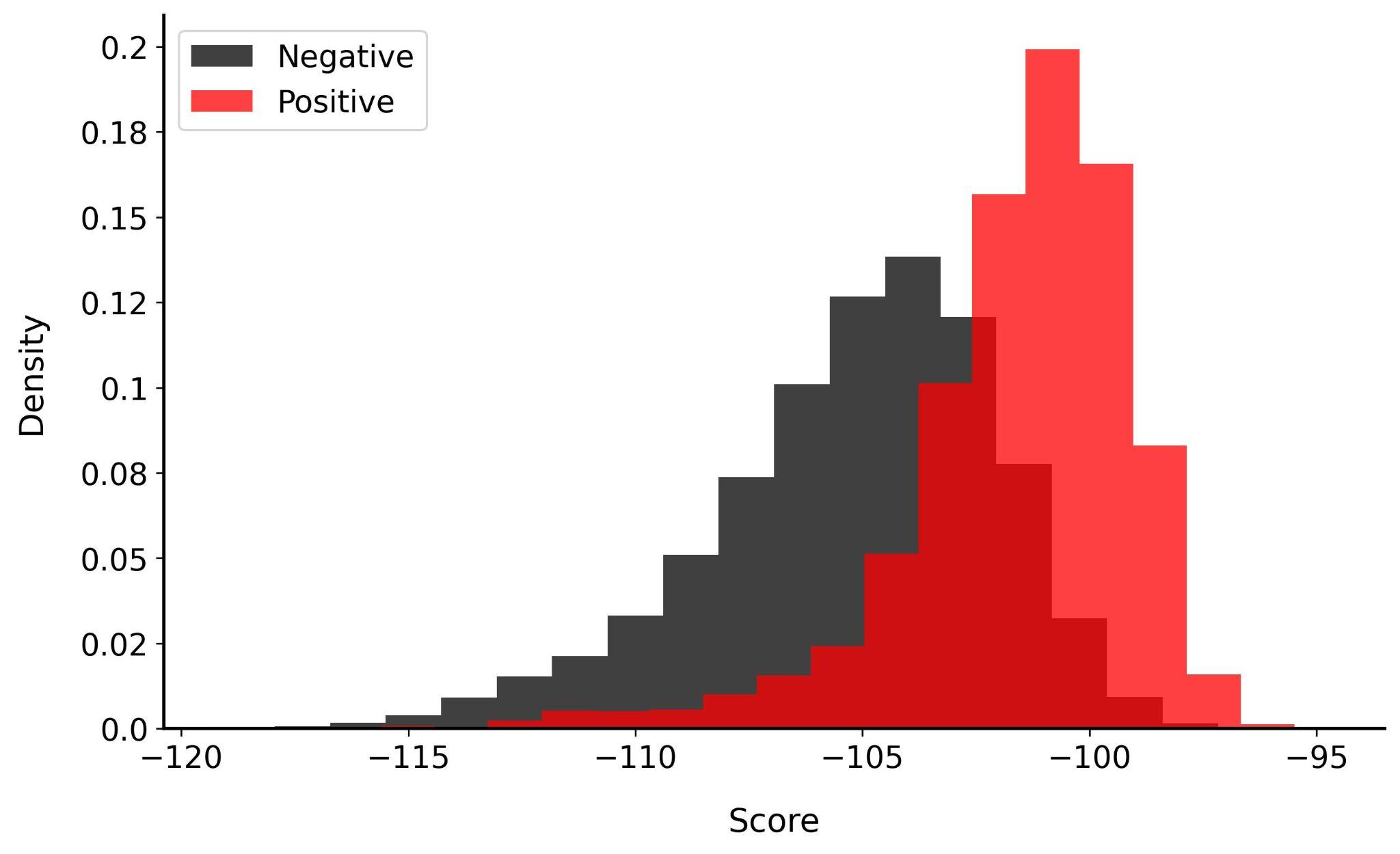 | |
| --- | --- |
| (b) **External Evaluation (Observed Drugs)**   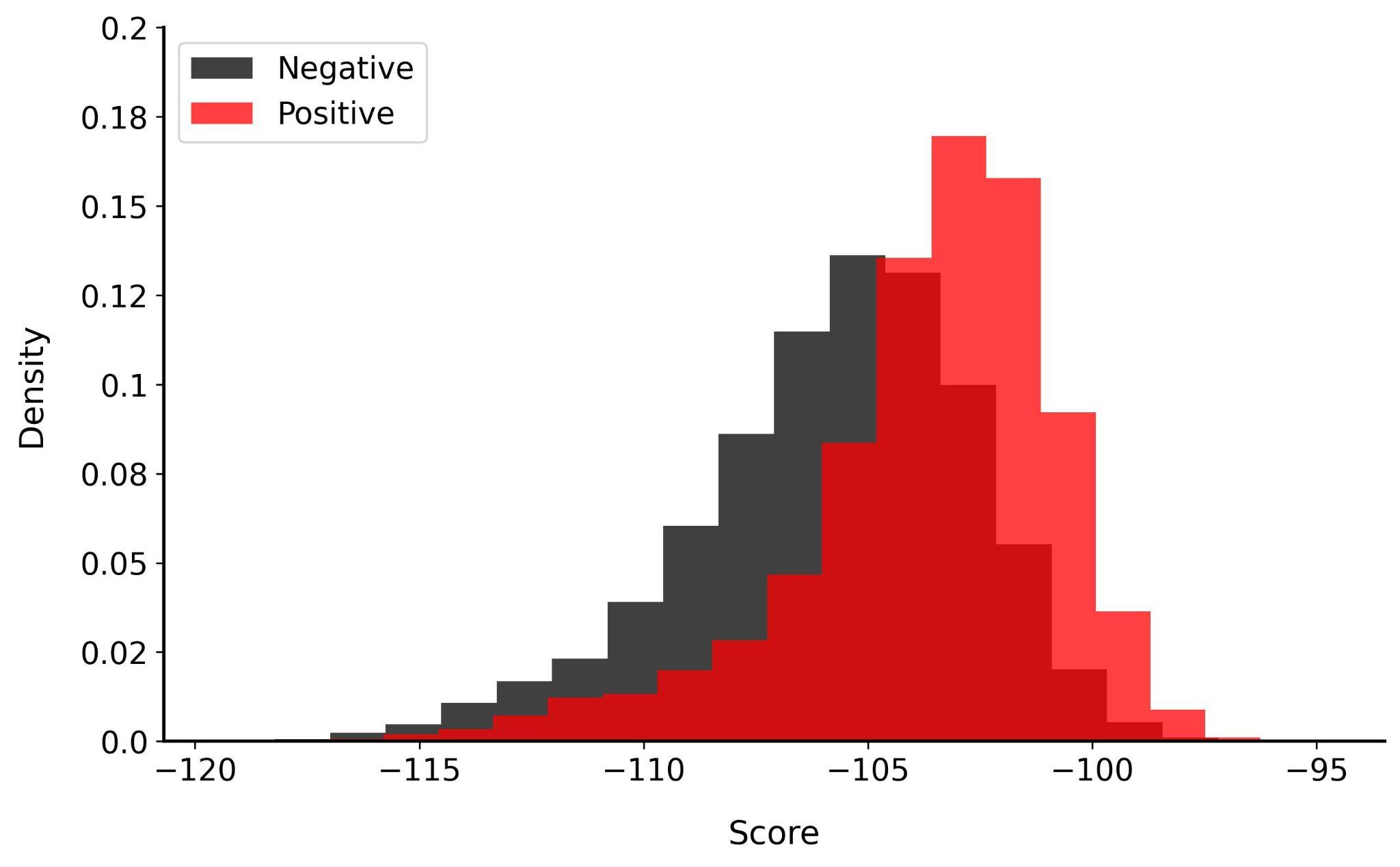 | **(c) External Evaluation (New Drugs)**  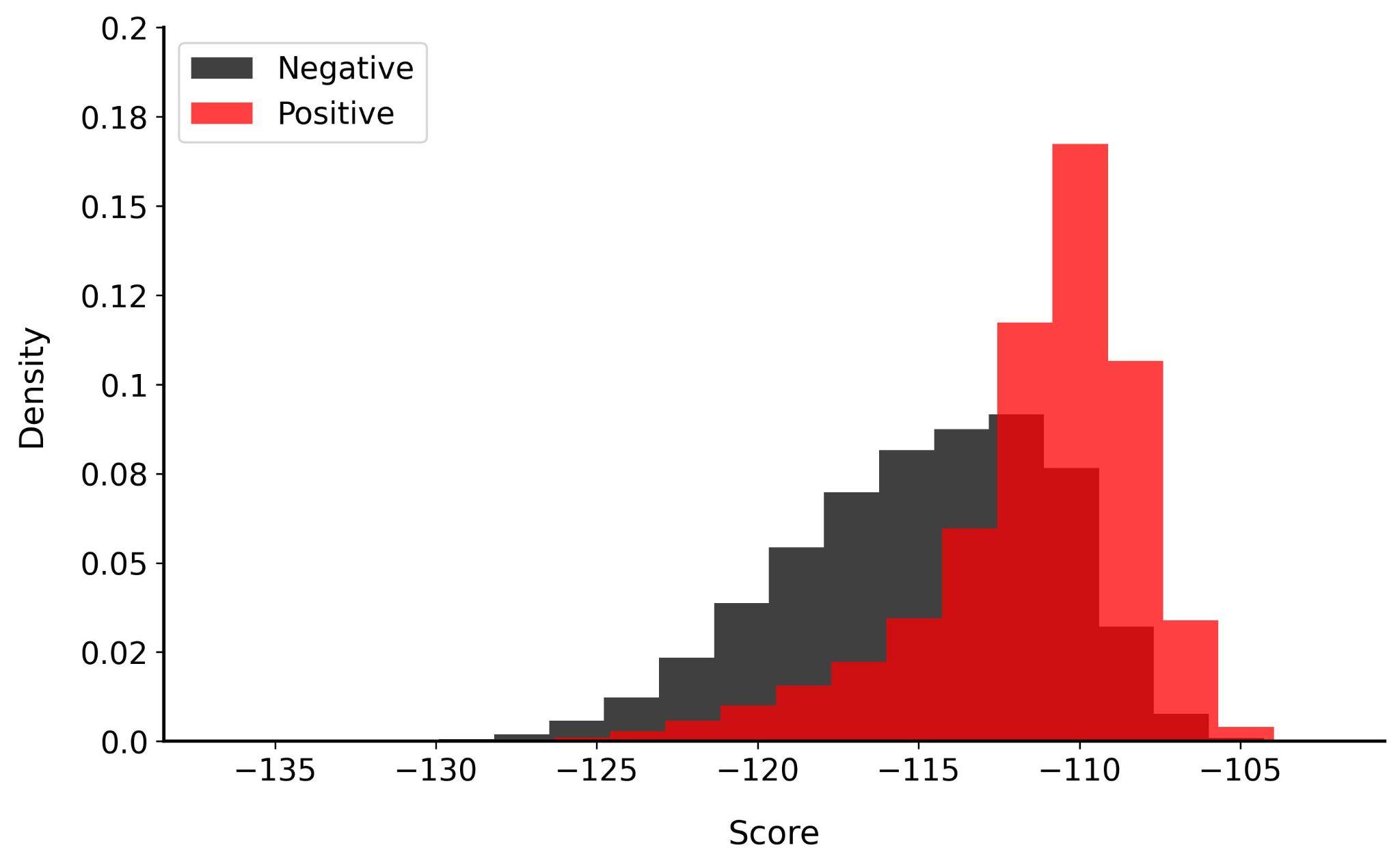 |

**Supplementary Fig. 3: The histogram of predicted TransE scores over positive and negative drug-SE edges in three evaluation scenarios.** The y-axis denotes the percentage of drug-SE edges with the TransE score.
